# Heat Exposure Among Adult Women in Rural Tamil Nadu, India

**DOI:** 10.1101/2022.11.07.22282037

**Authors:** Aniruddha Deshpande, Noah Scovronick, Thomas F. Clasen, Lance Waller, Vigneswari Aravindalochanan, Kalpana Balakrishnan, Naveen Puttaswamy, Jennifer Peel, Ajay Pillarisetti

## Abstract

Exposure to heat is associated with a substantial burden of disease and is an emerging issue in the context of climate change. Heat exposure is of particular concern in India – one of the world’s hotter countries and soon to be its most populous – where a large fraction of the population works outdoors, lives in dwellings that are thermally inefficient, and is unable to access cooling technologies. Despite these concerns, relatively little is known about personal heat exposure in India, particularly in rural areas. Here we leverage temperature data collected as part of a randomized controlled trial of cookstove replacement to describe personal temperature exposures of older adult women in rural Tamil Nadu. We also compare personal exposure measurements to the nearest ambient monitoring stations, as well as to commonly used modelled temperature data products. We find that temperatures differ across individuals in the same location on the same day – sometimes by more than 5 °C within the same hour – and that some individuals experience sharp increases in heat exposure in the early morning or evening, potentially a result of cooking with solid fuels. In comparisons with the personal exposure measurements, we find stronger correlations with the modelled products (R^2^ of ∼0.74) than with ambient monitors (R^2^ of ∼0.6). We did not find evidence of systematic biases, which indicates that adjusting for discrepancies between personal and ambient exposure estimates is not straightforward. This study indicates a need for improved heat exposure assessment in epidemiological and burden of disease studies in India.

## Introduction

Exposure to hot temperatures is a top environmental risk factor for global mortality.^1,2^ In 2019, an estimated 308,000 deaths were attributed to heat exposure^3^; this already substantial burden is expected to increase as the climate continues to warm.^4^ Heat is also associated with a substantial morbidity burden, as well as with reductions in labor productivity.^5-8^ Heat exposure is of particular concern in India one of the world’s hotter countries and soon to be its most populous^9^ – where a large fraction of the population works outdoors, lives in dwellings that are thermally inefficient, and is unable to access cooling technologies such as fans or air conditioners.^10^

Despite these concerns, relatively little is known about personal exposure to outdoor temperatures in India, particularly in rural areas. Ambient monitoring stations are sparse, and even where present may not accurately represent individual exposures, as people frequently move between indoor and outdoor environments, both in the sun and in the shade. Improving exposure assessment for temperature can enhance our understanding of the health effects of heat and cold by reducing potential biases and measurement error associated with ambient monitors and modeled products, which are commonly used in epidemiological and burden of disease studies.^1,11-13^ Personal measurements may also highlight opportunities for intervention by identifying high-exposure activities.

In this study, we leverage data collected as part of the Household Air Pollution Intervention Network (HAPIN) randomized controlled trial of cookstove replacement to describe personal temperature exposures of older adult women in rural Indian villages in the state of Tamil Nadu. In addition, we compare personal exposure measurements to the nearest identified ambient monitoring stations, as well as to two sources of modelled temperature data often used in health effects studies (due in part to the limited spatial coverage of the ambient monitoring network).^11,12^ Through these comparisons, we assess potential measurement error when using proxies for personal temperature exposure.

## Methods

### The HAPIN trial: overview, study site, and data collection

The HAPIN multi-country randomized controlled trial (RCT) is evaluating the effect of a liquefied petroleum gas stove and fuel intervention during pregnancy on birth weight, growth, and severe pneumonia in children and on blood pressure among older adult women (40 – 79 years of age). The trial’s research sites are in four diverse low- and middle-income settings: Guatemala, India, Rwanda, and Peru. The study began in 2017 and is ongoing. Details of the HAPIN trial have been published elsewhere^14,15^. The trial is registered with ClinicalTrials.gov (Identifier NCT029446282).

We focus exclusively on older adult women participants from the Indian site of the HAPIN trial, which consists of two districts, Villupuram and Nagapattinam, in Tamil Nadu (Figure 1). The hilly Villupuram study site is located at an altitude of approximately 800m above sea level, while the Nagapattinam site, a coastal area, is located at an average elevation between 10 and 50 m above the sea level (full details on the sites, and how they were selected, are in Sambandam et al., 2020^16^).

**Figure 1.**
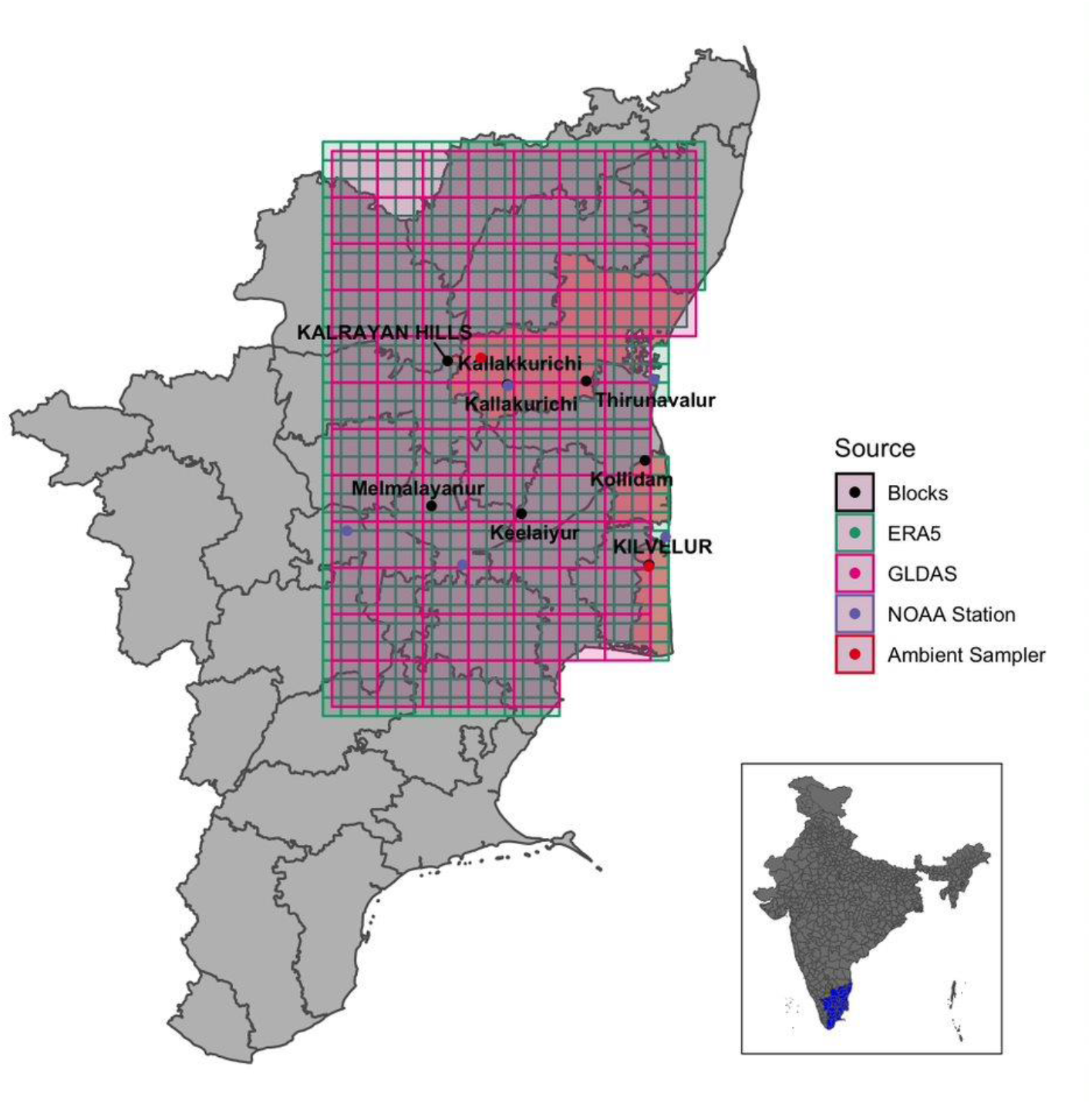
Map of the study villages and ambient monitors, overlaid with grids from the two modelled temperature products.^17^.

As part of the trial, participants are asked to wear a vest holding an Enhanced Children’s MicroPEM™ (ECM, RTI International, North Carolina, USA), a robust, lightweight, and validated nephelopmetric and gravimetric PM_2.5_ monitor. The ECM weighs approximately 150g and is capable of operating continuously for up to 48 h. These instruments additionally measure temperature and humidity to correct real-time estimates of air pollution levels; here, we take advantage of these measurements as reflective of temperatures experienced by participants as they move through space and time. Temperature is logged every 30 seconds. Participants were asked to wear the vest while awake during the day, and to hang the vest nearby when it is not being worn (such as while bathing or sleeping). The study sought to take 24-hr measurements on at least 3 occasions for each participant over the course of the 18-month HAPIN follow-up period.

#### HAPIN ambient monitors

In addition to personal exposure assessment, two ambient PM_2.5_ monitors (Metone E-Samplers, Grants Pass, Oregon, USA) were installed in the HAPIN study districts in Tamil Nadu to measure outdoor air pollution (Figure 1). They also log meteorological parameters, at 5-minute resolution. We use these monitors – which we refer to as ‘HAPIN ambient monitors’ – as one point of comparison with the personal monitors.

#### GHCN ambient monitoring stations

As a second point of comparison with the HAPIN personal monitors, we obtained contemporaneous temperature measurements from the nearest established ambient monitoring stations available from the archive of the Global Historical Climatology Network (GHCN), accessed from the US National Oceanographic and Atmospheric Administration’s National Centers for Environmental Information. The data is daily and was extracted using the R package *rnoaa*. The location of the stations relative to the study locations can be found in Figure 1.

#### Modelled temperature data

As a final point of comparison, we extracted contemporaneous temperature estimates from two modelled data products. The first is the ERA5-LAND product, which is a high-resolution (9km) reanalysis dataset based on the H-TESSEL land-surface model.^18^ The dataset provides hourly estimates at 2-meters above the land surface. ERA-5 data has been increasingly used in health effects studies.^1,13,19,20^ The second product is NASA’s GLDAS-2 product^21^, which provides temperature estimates every three hours at a spatial resolution of 0.25×0.25 degree, a scale more coarse than ERA-5 (Figure 1). GLDAS generates its estimates by fusing satellite- and ground-based observational data products, using advanced land surface modeling and data assimilation techniques.^21^

### Data analysis

First, we calculated descriptive statistics summarizing the personal exposure measurements by month and season, including mean temperature across all measurements, empirical distributions by month, and minima and maxima.

Next, we assessed the correlation between the personal exposures and corresponding estimates from alternate data sources. For comparison with temperatures measured by HAPIN ambient monitors, we matched all observations in each district to the closest corresponding station. To identify the closest GHCN station by Euclidean distance, we utilized the rnoaa R package. Finally, for comparison with the two modelled products (ERA5 and GLDAS), we used the GPS coordinates of each participant’s block of residence to assign the relevant grid square. As the different exposure sources provide measurements at varying temporal resolutions, correlations are based on daily average exposures.

In order to further summarize differences between the personal measurements and the alternate data sources, we produced Bland-Altman plots.^22^ Bland-Altman plots are a data visualization tool designed to characterize the agreement between two different data sources or measurement techniques, displaying the variance between the two measurements, the direction of any bias, and if/how the bias changes along the exposure (temperature) distribution. The Bland-Altmann analyses provide associations between bias and the average temperature for the measurement data being compared. This not only allows us to assess the strength of agreement between two sources, similar to a correlation coefficient, but also how agreement varies across the temperature distribution observed. All Bland-Altmann analyses were generated relative to the personal measurements, again using daily averages for consistency across data sources.

All analyses were performed in R version 4.1.3 (R Foundation for Statistical Computing, Vienna, Austria).

### Ethics

The study protocol has been reviewed and approved by institutional review boards (IRBs) and Ethics Committees at Emory University (00089799), Johns Hopkins University (00007403), Sri Ramachandra Institute of Higher Education and Research (IEC-N1/16/JUL/54/49), the Indian Council of Medical Research – Health Ministry Screening Committee (5/8/4-30/(Env)/Indo-US/2016-NCD-I), Universidad del Valle de Guatemala (146-08-2016), Guatemalan Ministry of Health National Ethics Committee (11-2016), Asociación Benefica PRISMA (CE2981.17), the London School of Hygiene and Tropical Medicine (11664-5), the Rwandan National Ethics Committee (No.357/RNEC/2018), and Washington University in St.Louis (201611159). The study has been registered with ClinicalTrials.gov (Identifier NCT02944682).

## Results

### Personal measurements

A total of 612 measurements (approximately 1.7 million datapoints) were recorded from 104 different participants. Participants were measured on average 5.8 times (SD 2.2, range 1 – 11). The first measurement was on 13 June 2018 and the last one available in this dataset was recorded on 28 June 2021. The average age was 49 years (SD = 3.9 years), most participants received little formal education, and most work in agriculture (Table 1). House construction was a mix of traditional (e.g. thatch/ceramic/mud) and modern (e.g. concrete) materials and no household had air conditioning.

**Table 1.** Baseline characteristics of study population.

| Characteristics |  | India (N=104) |  |
| --- | --- | --- | --- |
| Participant characteristics |  |  |  |
| Age at screening |  |  |  |
| Mean (SD) |  | 49.0 | (6.5) |
| Highest level of education |  |  |  |
| No formal education or primary school incomplete |  | 99 | (95%) |
| Primary school complete |  | 5 | (5%) |
| Main occupation |  |  |  |
| Agriculture |  | 87 | (84%) |
| Household |  | 10 | (10%) |
| Unemployed |  | 7 | (7%) |
| Other |  | 9 | (9%) |
| Household characteristics |  |  |  |
| Household size |  |  |  |
| Mean (SD) |  | 4.4 | (1.3) |
| Roof type in main home |  |  |  |
| Thatch |  | 28 | (27%) |
| Concrete |  | 28 | (27%) |
| Ceramic/Fired Tile |  | 25 | (24%) |
| Other |  | 23 | (22%) |
| Wall type in main home |  |  |  |
| Concrete |  | 55 | (53%) |
| Mud |  | 39 | (38%) |
| Other |  | 10 | (10%) |
| Floor type in main home<sup>1</sup> |  |  |  |
| Concrete |  | 58 | (56%) |
| Mud |  | 42 | (40%) |
| Other |  | 6 | (6%) |
| Air cooler/air conditioner |  |  |  |
| No |  | 104 | (100%) |
| Time to take to go get water and come back (minutes) |  |  |  |
| Mean (SD) |  | 34.2 | (32.9) |
| Categorical household food insecurity<sup>2</sup> |  |  |  |
| None (0) |  | 83 | (80%) |
| Mild (1-3) |  | 17 | (16%) |
| Moderate/Severe (4-8) |  | 4 | (4%) |
| Baseline Exposure |  |  |  |
| Primary Fuel type |  |  |  |
| Wood |  | 104 | (100%) |
| Primary heating source |  |  |  |
| Do not use heating |  | 91 | (88%) |
| Traditional cookstove/Three-stone fire |  | 11 | (11%) |
| Other |  | 2 | (2%) |

<sup>1</sup>Multiple materials may be reported for the same household, so households may appear more than once
<sup>2</sup>The Food Insecurity Experience Scale - Developed by the Food and Agriculture Organization of the United Nations, http://www.fao.org/3/as583e/as583e.pdf

Density functions of 30 second personal temperature exposure on the HAPIN participants by month for each of three seasons – winter, summer, and monsoon – are reported in Figure 2. The overall average temperature exposure was 28.1 °C (SD 3.2), with seasonal differences; average exposure was 26.2 °C in winter (SD 2.9), 30.3 °C (SD 2.9) in summer, and 28.1 °C (SD 3.0) during the monsoon.

**Figure 2.**
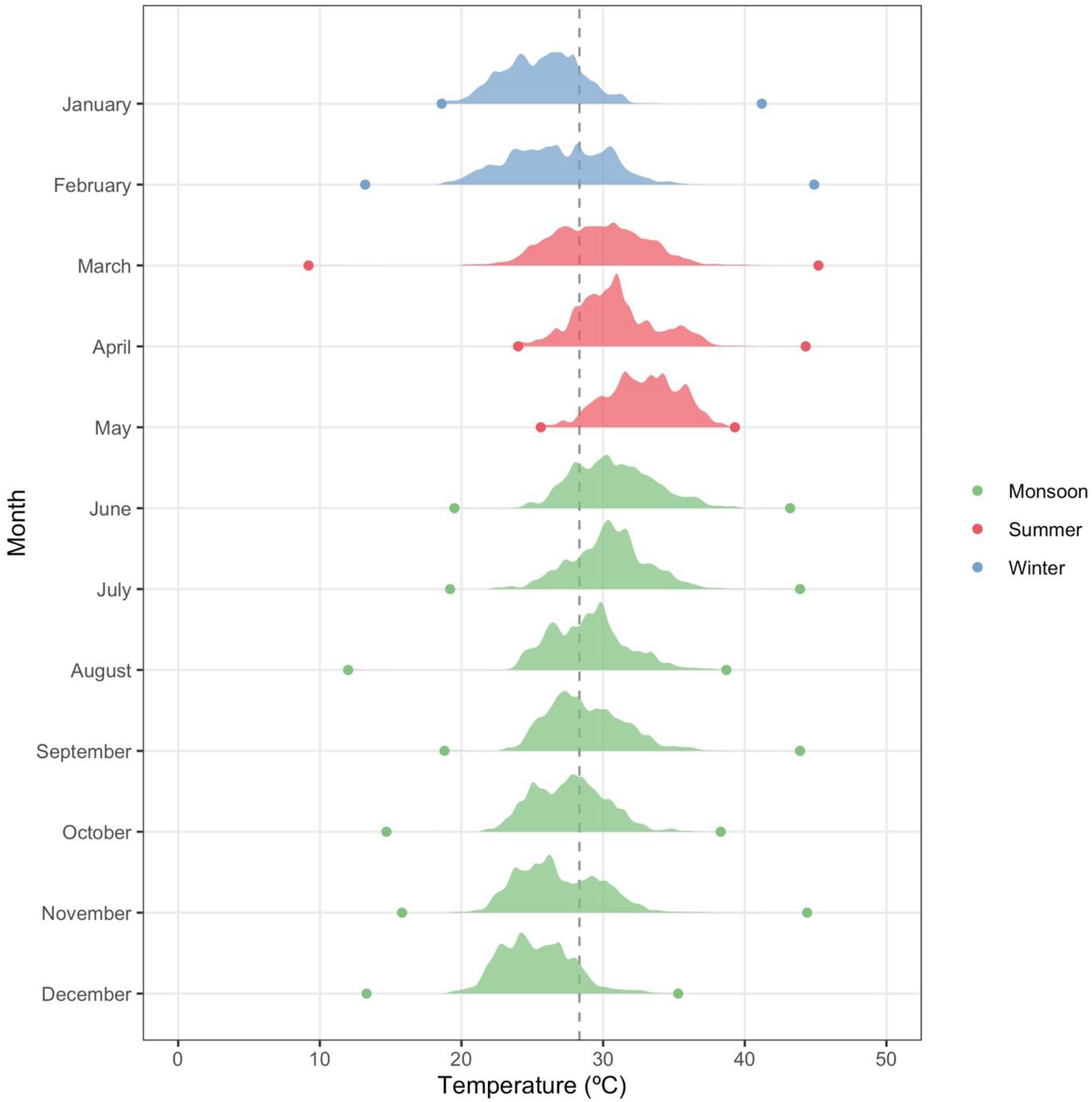
Density plots of personal exposure by month during the 2018-2021 study period. The vertical line indicates the average temperature over the study period. Dots are monthly average minimum and maximum values.

Figure 3 presents hourly personal exposures from multiple individuals for the same 24-hr period, starting at 8:00 am on 25^th^ November 2019. Two heat exposure-related features of interest in the study area are illustrated in this figure. First, temperatures differ across individuals on the same day, sometimes by more than 5 °C within the same hour. Second, the data suggests that some individuals experience sharp increases in heat exposure in early morning or late afternoon/evening, potentially a result of cooking with solid fuels and, thus, proximity to stoves or other combustion sources (for example, for heating). During these times, heat exposure for an individual can vary by several degrees within an hour.

**Figure 3.**
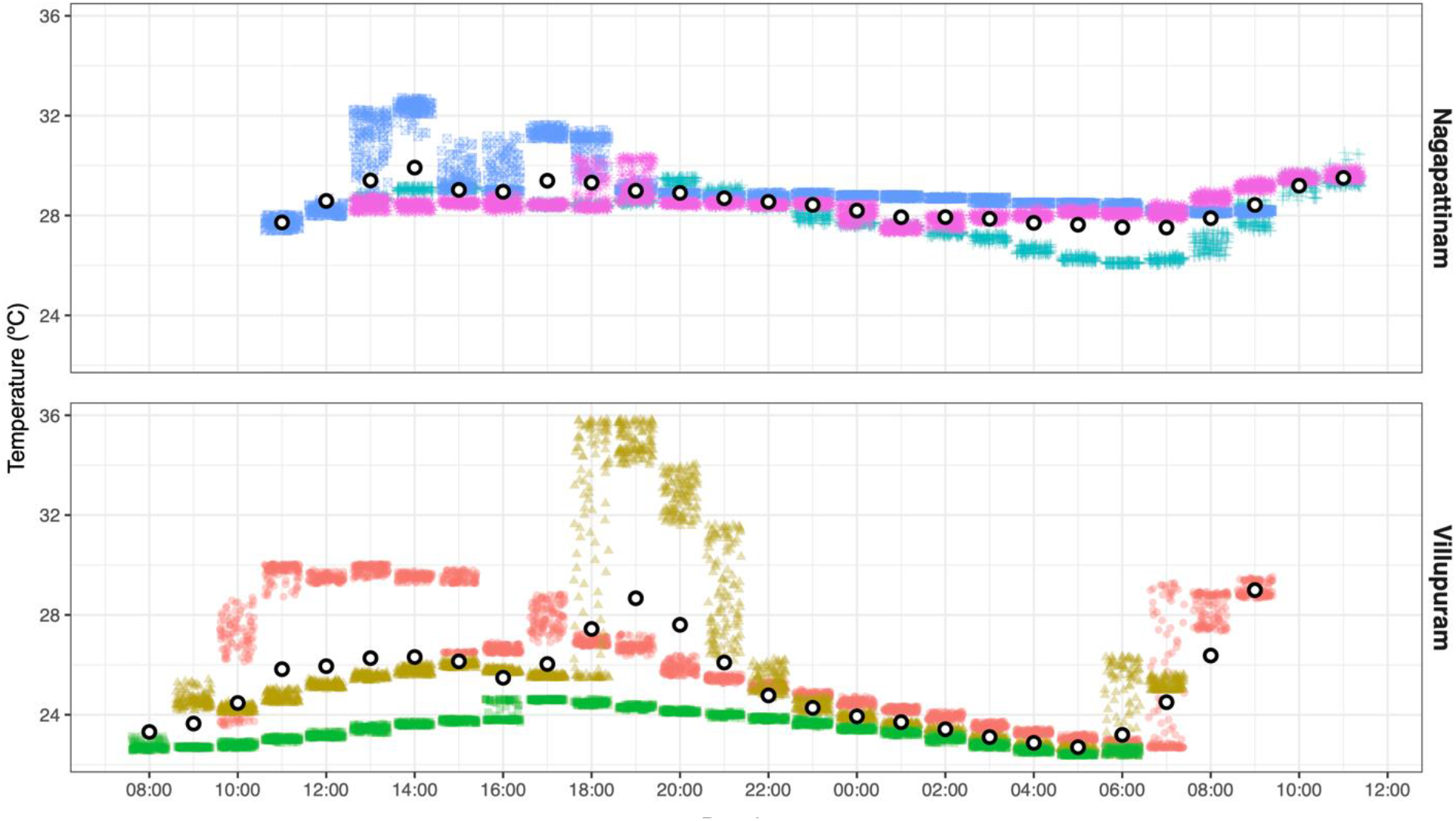
Personal exposure of six individuals (three in each district) from 8:00 am on 11/25/2019 to 8:00 am on 11/26/2019. Each individual is represented by a unique color-shape combination and each small colored shape represents a single measurement. White points with a black outline are average hourly measurements in each village.

### Comparison of exposure sources

Summary statistics overall and by season for the different temperature sources are reported in Table 2. In general, the personal measurements tend to be intermediate between the lower temperatures reported by the modelled products and the higher temperatures reported by the ambient monitors.

**Table 2.** Daily summary statistics by data source (overall and by season).

| Data Source | All Seasons |  | Monsoon |  | Summer |  | Winter |  |
| --- | --- | --- | --- | --- | --- | --- | --- | --- |
|  | Mean (SD) | Range | Mean (SD) | Range | Mean (SD) | Range | Mean (SD) | Range |
| HAPIN personal | 28.1 (3.2) | 19.6-36.3 | 28.1 (3.0) | 20.1-36.3 | 30.3 (2.9) | 22.4-36.3 | 26.2 (2.9) | 19.6-32.8 |
| HAPIN ambient | 28.5 (3.3) | 19.1-37.6 | 28.3 (3.2) | 22.2-37.5 | 31.1 (3.8) | 19.9-37.6 | 27.6 (2.6) | 19.1-32.3 |
| GHCN ambient | 29.5 (2.6) | 24.2-35.9 | 29.9 (2.5) | 24.2-34.9 | 31.1 (2.1) | 27.4-35.9 | 27.0 (1.3) | 24.3-31.8 |
| ERA5 | 25.7 (2.9) | 19.5-34.2 | 25.5 (2.8) | 19.5-34.2 | 27.9 (2.3) | 22.2-34.2 | 24.1 (2.2) | 19.5-27.9 |
| GLDAS | 25.6 (3.4) | 18.1-35.9 | 25.5 (3.4) | 18.1-33.7 | 28.3 (2.8) | 22.2-35.9 | 23.4 (2.2) | 18.4-27.5 |

Scatterplots and correlations between personal exposures and alternate data sources are shown in Figure 4. Over the full study period, there were stronger correlations with the modelled products (R^2^ for both = 0.74) than with the semi-local ambient monitors (R^2^ for both ∼ 0.6). Correlations varied across seasons, but were uniformly highest in the monsoon season, and, with the exception of the GHCN data, was lowest in the winter.

**Figure 4.**
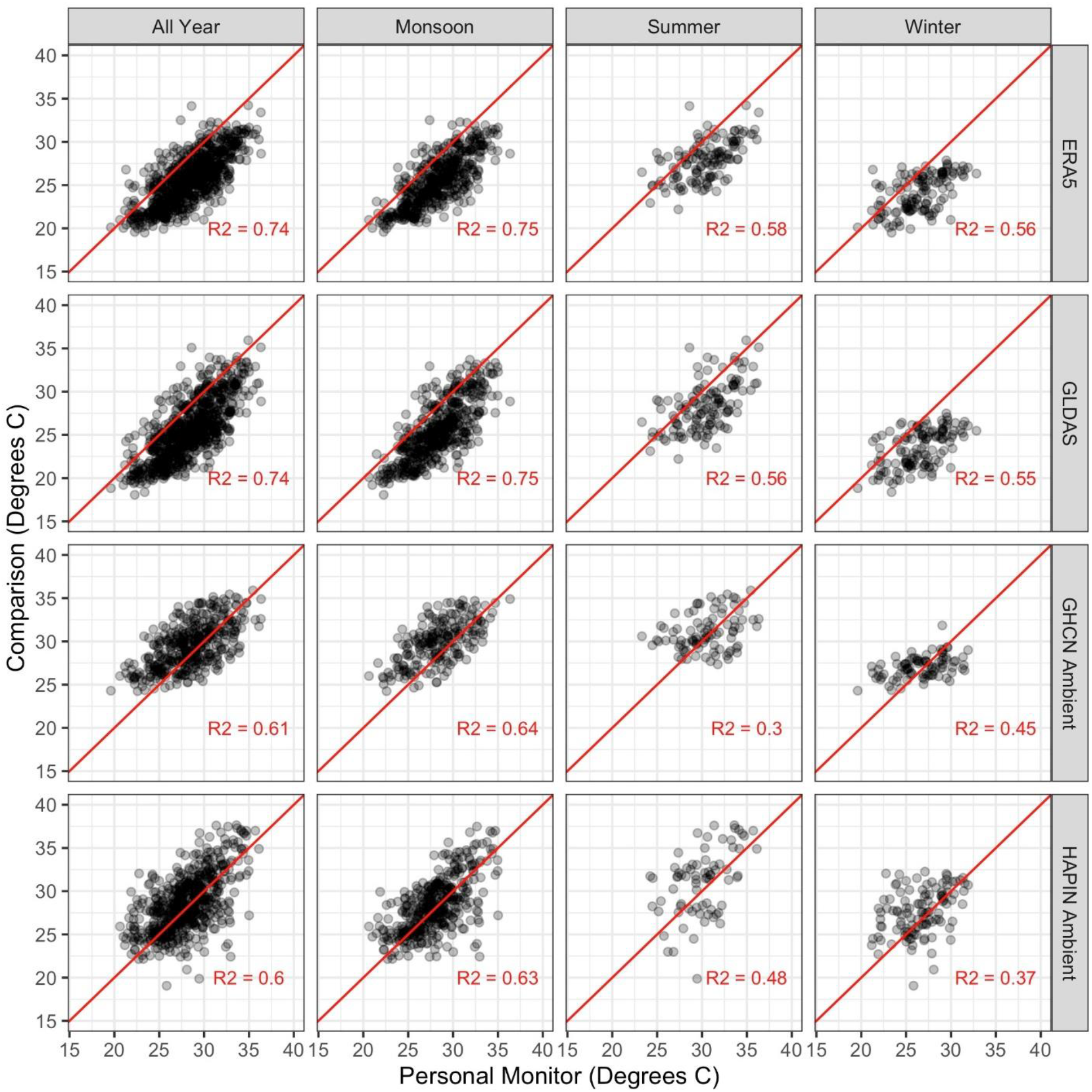
Scatterplots and simple correlations comparing personal exposures with ambient monitors and modelled products. Red lines are 1:1 lines; points represent daily average values.

The Bland-Altmann plots (Figure 5) indicate that the mean bias for both the ERA5-Land and GLDAS products was negative for the majority of measurements in all seasons. In contrast, the distribution for the GHCN and HAPIN ambient sampler data is centered closer to zero, with a relatively even number of measurements with positive and negative biases. In the all-year analyses for all sources, slopes were < +/-0.25, indicating that the bias remained similar across the temperature distribution. Confidence bands were wider for the monitors compared to the modelled products, indicating more error.

**Figure 5.**
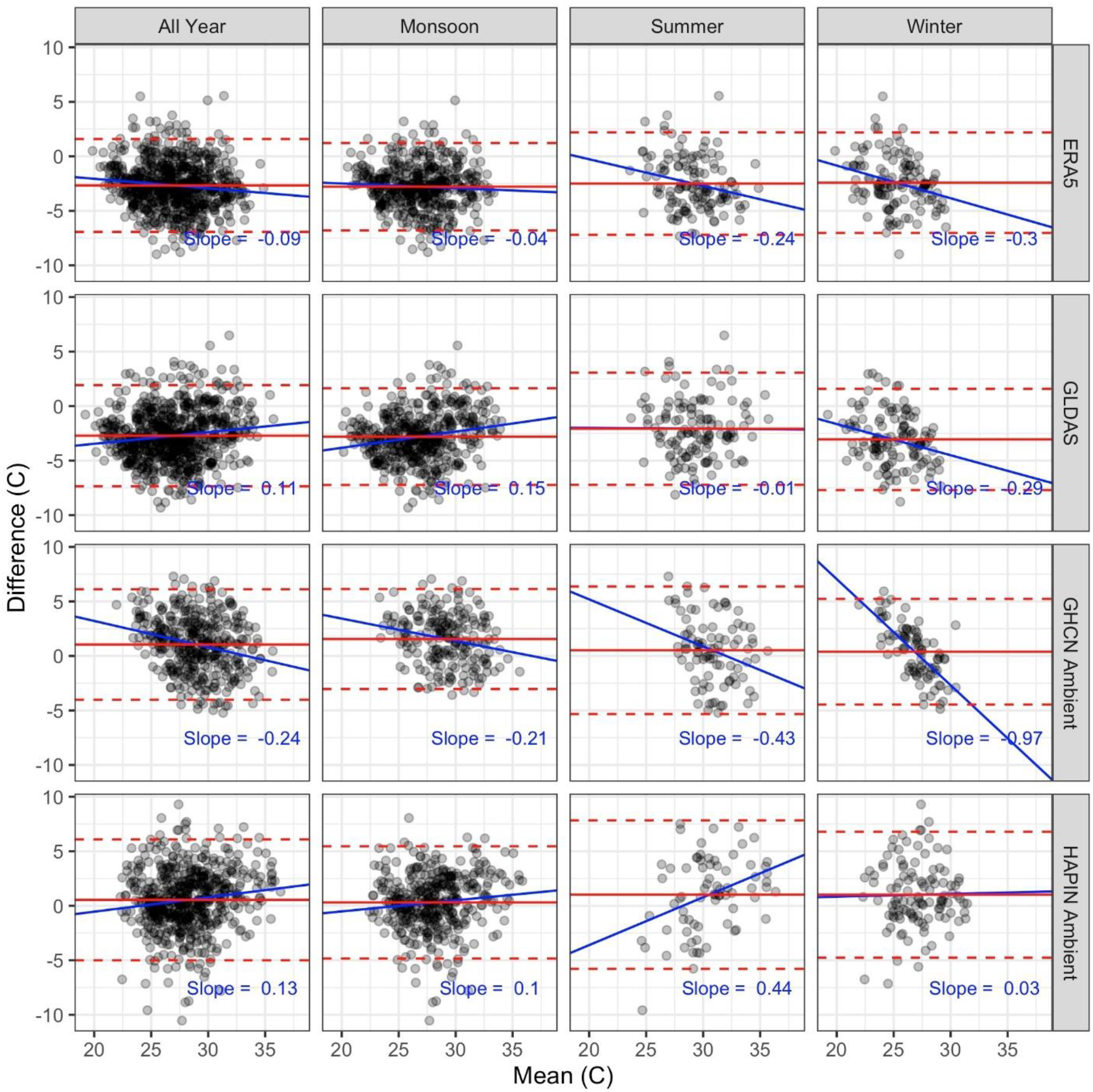
Bland-Altman plots comparing personal exposures with ambient monitors and modelled products. Blue sold lines are best-fit regression lines displaying the relationship between bias and mean changes in daily temperature. Dashed red lines are 95% Wald confidence intervals; solid red lines are mean values.

## DISCUSSION

We have presented the results of opportunistic monitoring of personal temperature exposures in Tamil Nadu, India, that were collected as part of a large-scale household air pollution intervention study. To our knowledge, this is one of very few studies of its kind in India – a country highly vulnerable to climate change and extreme heat – and the first conducted in multiple rural districts. We find that personal measurements follow expected seasonal trends but that even within a season or month, individuals experience a wide range of temperature exposure. The average exposure across the study period was 28.3 °C, but in all months the study participants experienced many exposures above 30-35 °C; days with exposures above 40 °C occurred in most months. We also found that differences of several degrees may be evident across individuals in the same district even within the same hour of the same day, and that individuals themselves may have highly variable exposures within a short period of time. In some individuals, the daily pattern of heat exposure is suggestive of cooking and/or heating with solid fuels and other behaviors that likely impact exposure.

A previous study in peri-urban Telangana, India compared personal measurements of temperature opportunistically collected from a similar monitor to the one employed in HAPIN with ambient measurements among a population of 50 participants.^23^ They noted limited agreement between personal and ambient samplers and suggest that additional factors – like altitude and demographic data may help explain the discordance between the monitoring types. To the best of our knowledge, that study did not investigate relationships between modeled ambient temperature products and personal exposure, as we did here.

We also compared our data from personal monitors with contemporaneous data from study and government ambient monitors and two gridded meteorological products. In general, the modelled products performed best, having a higher correlation with the personal measurements and smaller mean errors, as shown in the Bland-Altman analyses. This information may be relevant in the choice of exposure data when conducting observational studies on the relationship between temperature and health (or non-health) outcomes. Nevertheless, differences were often ≥ 3-5 degrees in either direction, which may be problematic for the design of interventions to protect against extreme heat. The finding that there seemed to be no systematic relationship between the personal measurements and the alternative data sources indicates that adjusting for the discrepancies is not straightforward. The overall implication is that epidemiological studies based on existing options for exposure assessment may introduce exposure misclassification and therefore produce imprecise and/or inaccurate health effects estimates. Such misclassification could also affect burden of disease calculations.

This study has several important limitations and raises the need for additional research. One key limitation is that we only present data for older adult women, which may not be representative of the study population at large. Even within the population of older adult women, more measurements would enhance the robustness of the results, particularly with respect to potential seasonality in correlations with the ambient monitors and modelled products. We also emphasize that the performance of these alternative sources in Tamil Nadu does not necessarily hold in other study locations, particularly those with more (and closer) monitoring stations. However, for many rural locations in low- and middle-income countries, we expect that similar discrepancies may be apparent.

Future research can replicate these results with more data, in other locations, by exploring the drivers of differences between data sources, and by explicitly analyzing how potential biases may influence epidemiological or econometric studies on the consequences of heat exposure. We believe there are many opportunities to leverage existing data to answer these and other questions. Real-time particulate matter sensors have been used in hundreds of settings in dozens of contexts around the world to assess exposure to household air pollution arising from the use of solid fuels for cooking and heating. Because these real-time particulate matter monitors must measure temperature (and often also measure humidity), there is potentially a large amount of existing data that can be analyzed to characterize and describe heat exposures across a swath of a typically unmonitored population. Furthermore, given the proliferation of these types of sensors around the globe – as part of primarily urban low-cost air monitoring networks, like the Purple Air network – there may be utility in assessing the information they provide on heat and humidity at a finer geographic and temporal scale. Such opportunistic monitoring may enable more advanced epidemiological analyses and provide better estimation of personal temperature exposure.

## Data Availability

All data produced in the present study are available upon reasonable request to the authors

